# Knowledge and stigma of autism spectrum disorders in Chinese university students in the context of inclusive education

**DOI:** 10.1101/2025.02.27.25322274

**Authors:** Jinping Hu, Pengwei Fu, Shiying Qiao, Xinai Yan

## Abstract

This study investigated Chinese university students’ autism spectrum disorder knowledge and stigmatizing attitudes using the cross-culturally validated Chinese ASK-Q scale through an online survey of 2,081 students across 25 provinces. Independent samples t-tests and ANOVA revealed that females, upper-grade students, normal-education students, special education majors, and those enrolled in inclusive education courses exhibited significantly higher ASD knowledge and lower stigma endorsement. Interaction with autistic individuals was significantly associated with enhanced knowledge and reduced stigma. The findings suggest that universities should enhance the accessibility of special education courses, foster opportunities for future educators to engage with autistic communities, and strengthen inclusive education competencies to mitigate stigma and improve professional preparedness.

## Introduction

Inclusive education represents the trajectory of global special education development, embodying the modern society’s pursuit of educational equity[1–4]. In recent years, the Chinese government has vigorously promoted inclusive education, and with the launch of the “Fourteenth Five-Year Plan for Special Education Development and Enhancement Action Plan,” it has become a core strategy for improving the quality of special education. As more children with special needs enter general education schools, general education teachers face new role demands. They are required not only to possess general educational knowledge but also to master skills related to inclusive and special education, that is, inclusive education literacy, to provide effective support [5,6]. Constructing a teaching workforce with professional inclusive education capabilities is a critical task, and the cultivation of general teachers’ inclusive education literacy during the pre-service education stage is especially important [7–11]. The Chinese Ministry of Educatio’s 2021 standards for normal student education explicitly included the concept and skills of inclusive education into the pre-service teacher education standards [12].

Autism Spectrum Disorder (ASD) is a type of neurodevelopmental disorder primarily occurring in childhood. In recent years, the global prevalence of ASD has risen rapidly. The Centers for Disease Control and Prevention (CDC) estimated in 2023 that one in every 36 children is diagnosed with ASD [13]. In China, there are over two million autistic children, with approximately 150,000 new cases each year [14]. With the increase in diagnosed cases, how to provide suitable educational environments for autistic children has become an important issue. The promotion of inclusive education brings diverse curriculum options, skill enhancement, and social interaction benefits to autistic children[15], and also helps to reduce stereotyping and stigmatization [16–18].

The concept of “stigma”was introduced by Goffman, referring to the social exclusion suffered due to negative labeling, with autism often being perceived as a “hidden stigmatized identity,” leading to social exclusion [19,20]. Research indicates that the public’s understanding of ASD is insufficient [21,22], particularly among university students, which affects societal inclusivity towards ASD [23]. Enhancing awareness of autism has become one of the key goals of global mental health [24–26]. Surveys show that university students have limited understanding of ASD[7,27,28], with confusion or misconceptions [29]. Universities, as cradles for training inclusive education talent [30], as well as future drivers of inclusive education, hold a critical position for normal-education students’ cognition[31,32]. If normal-education students possess knowledge and skills related to children with special needs, it will aid them in becoming competent inclusive education teachers [33].

Conducting surveys on the knowledge of autism and stigmatizing attitudes among university students serves to discern the varying levels of awareness regarding autism spectrum disorder across a spectrum of student demographics. Such surveys inform the development of tailored educational policies that aim to foster a scientifically grounded understanding of autism among students, thereby elevating their consciousness towards embracing diversity within communities. In China’s higher education system, university students are categorized by their major into normal-education students and non-normal-education students. The subjects of this study include both normal-education students and non-normal-education students, with normal-education students further divided into those majoring in special education and those majoring in non-special education. Some universities offer “Inclusive Education”courses for normal-education students. Thus, the purposes of this study are to:

1. A comprehensive examination of the ASD knowledge and stigma of among Chinese university students with different demographic variables.
2. To test whether normal-education students have a higher level of ASD knowledge and a lower level of stigma than non-normal-education students. Within the normal-education student category, do special education majors or those who have taken inclusive education courses have a higher level of ASD knowledge and a lower level of stigma?

The specific hypothesis of this study is that normal-education students have more knowledge of ASD and a lower stigma identification than non-normal-education students. Among normal-education students, special education majors have the highest level of ASD knowledge and the lowest level of stigma, followed by normal-education students who have taken inclusive education courses, who have a higher level of ASD knowledge and a lower level of stigma.

## Methods

The Autism Stigma and Knowledge Questionnaire (ASK-Q) was developed to facilitate cross-cultural comparisons of ASD knowledge. During its development phase, international researchers evaluated the items within the ASK-Q to ensure its cross-cultural validity, thereby reducing the need for future cultural adaptations[34]. The questionnaire has demonstrated high internal consistency (Cronbach’s Alpha = 0.88)[35]. This study employed the Chinese version of the questionnaire, and the results indicated that the meticulously adapted Chinese ASK-Q performed well in a large national sample, validating ASK-Q as a tool for assessing ASD knowledge in a cross-cultural context [36]. Additionally, the ASK-Q exhibited acceptable internal consistency in the current sample, with an overall alpha coefficient of 0.773.

Procedures Following approval from the university’s review board, an online survey was created using Questionnaire Star (https://www.wjx.cn). The questionnaire was formatted into an online link and disseminated through WeChat or QQ social groups. A uniform informed consent description is provided in the questionnaire, which describes the purpose and significance of the study, as well as the completion method and precautions. Participants agree to participate in the survey by clicking on the link and answering the questions. All surveys are conducted anonymously. Recruitment for the study began on April 1 and ended on May 1, 2024. The questionnaire was divided into two sections: the first part collected demographic information, the specifics of which are presented in Table 1.

**Table 1.**
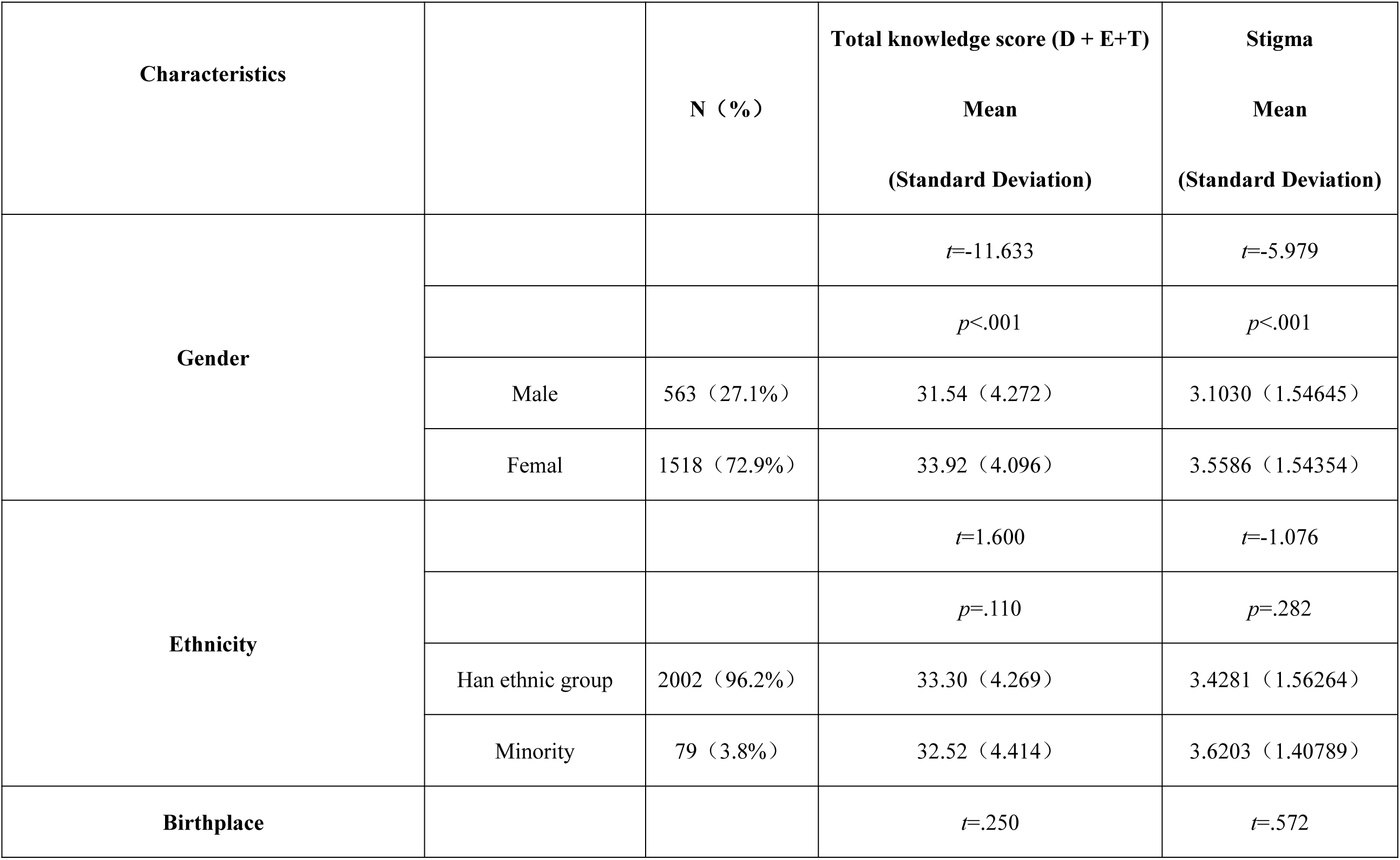
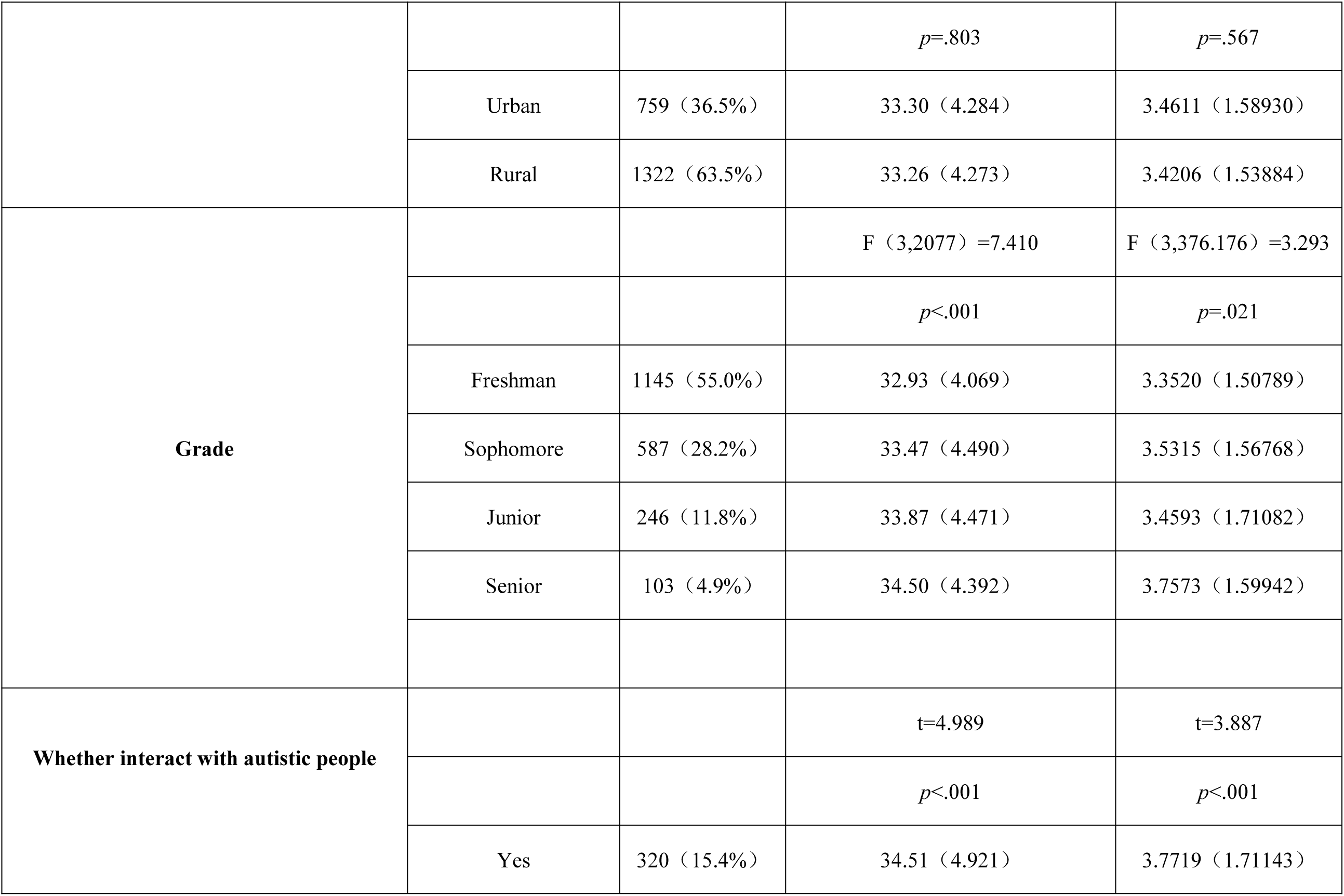
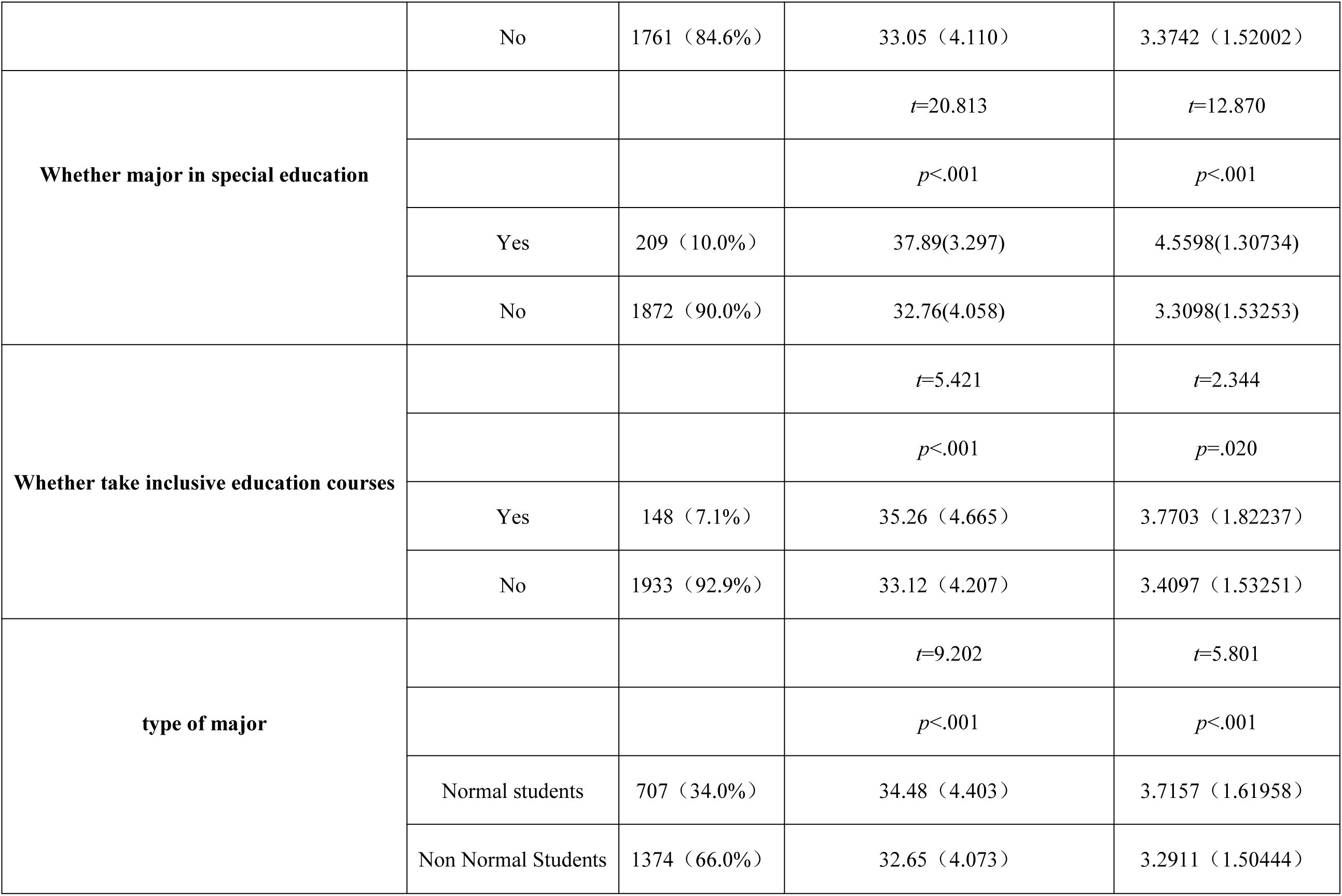
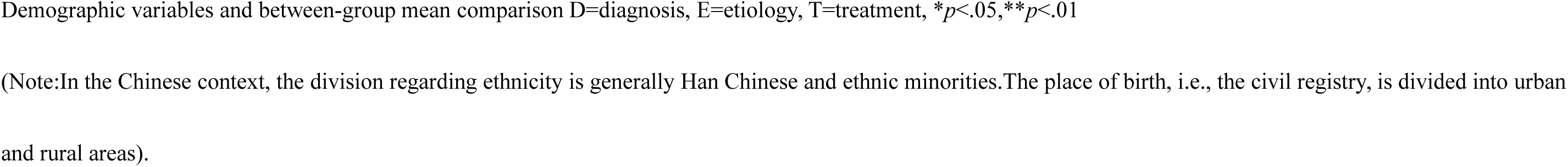
Demographic variables and comparison of means between groups.

The second part consisted of the 49-item ASK-Q questionnaire. The ASK-Q used a dichotomous scoring system, with options of “correct” or “incorrect.” One general knowledge question about ASD (“I have heard of autism”) was excluded from the scoring as it could not be judged as correct or incorrect. Therefore, the questionnaire comprised 48 items, with higher total scores indicating greater understanding of ASD, and higher stigma scores indicating lower stigma endorsement. To prevent random responses and ensure the quality of the questionnaire, the questionnaire included items with specified options, such as “Please select 2 for this question” to test if respondents were answering carefully. Any participant who answered these items incorrectly was considered to have completed the questionnaire invalidly, and their data were excluded. A total of 2564 questionnaires were collected, with 483 deemed invalid and excluded, resulting in 2081valid questionnaires, yielding an effective response rate of 81.2%.The screened data was exported to an Excel table and finally imported into SPSS 27.0 software for further data analysis.

## Results

### Comparison of different demographic variables

Independent sample t-tests and ANOVA analyses revealed significant effects of gender on the knowledge of ASD and stigma scores among university students (*p*<0.001). Female students scored higher in ASD knowledge (M=33.92, SD=4.096) and stigma (M=3.5586, SD=1.54354) compared to male students (ASD knowledge: M=31.54, SD=4.272; stigma: M=3.1030, SD=1.54645), indicating that females possessed greater knowledge of ASD and endorsed lower levels of stigma. Ethnicity and place of birth did not influence the scores. A positive correlation was found between grade level and ASD knowledge scores (*p*<0.001), with senior students achieving the highest scores and the lowest stigma endorsement. University students who had interacted with the autistic people had higher scores (ASD knowledge: M=34.51, SD=4.921; stigma: M=3.7719, SD=1.71143) than those who had not (ASD knowledge: M=33.05, SD=4.110; stigma: M=3.3742, SD=1.52002), indicating that interaction experience is associated with increased knowledge of ASD and reduced stigma endorsement.

### Comparative analysis of University students with different academic backgrounds

Normal-education students outperformed non-normal-education students in both ASD knowledge (M=34.48, SD=4.403 vs. M=32.65, SD=4.073, *p*<0.001) and stigma scores (M=3.7157, SD=1.61958 vs. M=3.2911, SD=1.50444, *p*<0.001), indicating superior ASD knowledge and lower stigma endorsement among normal-education students.

Special education majors scored significantly higher in ASD knowledge (M=37.89, SD=3.297, *p*<0.001) and stigma (M=4.5598, SD=1.30734, *p*<0.001) compared to non-special education majors, suggesting greater ASD knowledge and lower stigma endorsement among special education students.

Students with inclusive education courses scored higher in ASD knowledge (M=35.26, SD=4.665, *p*<0.001) and stigma (M=3.7703, SD=1.82237, *p*=0.020) than those without, indicating that inclusive education course takers have greater ASD knowledge and lower autism stigma endorsement.

### Sources of ASD knowledge among University students

As shown in Figure1,the internet (65%)and TV/movies (64%) were the most common sources of ASD knowledge, followed by social media (48%), news reports (41%), personal experience (39%), university courses (36%), doctors or other medical professionals (21%), research articles (18%), and club activities (15%).

**Fig 1.**
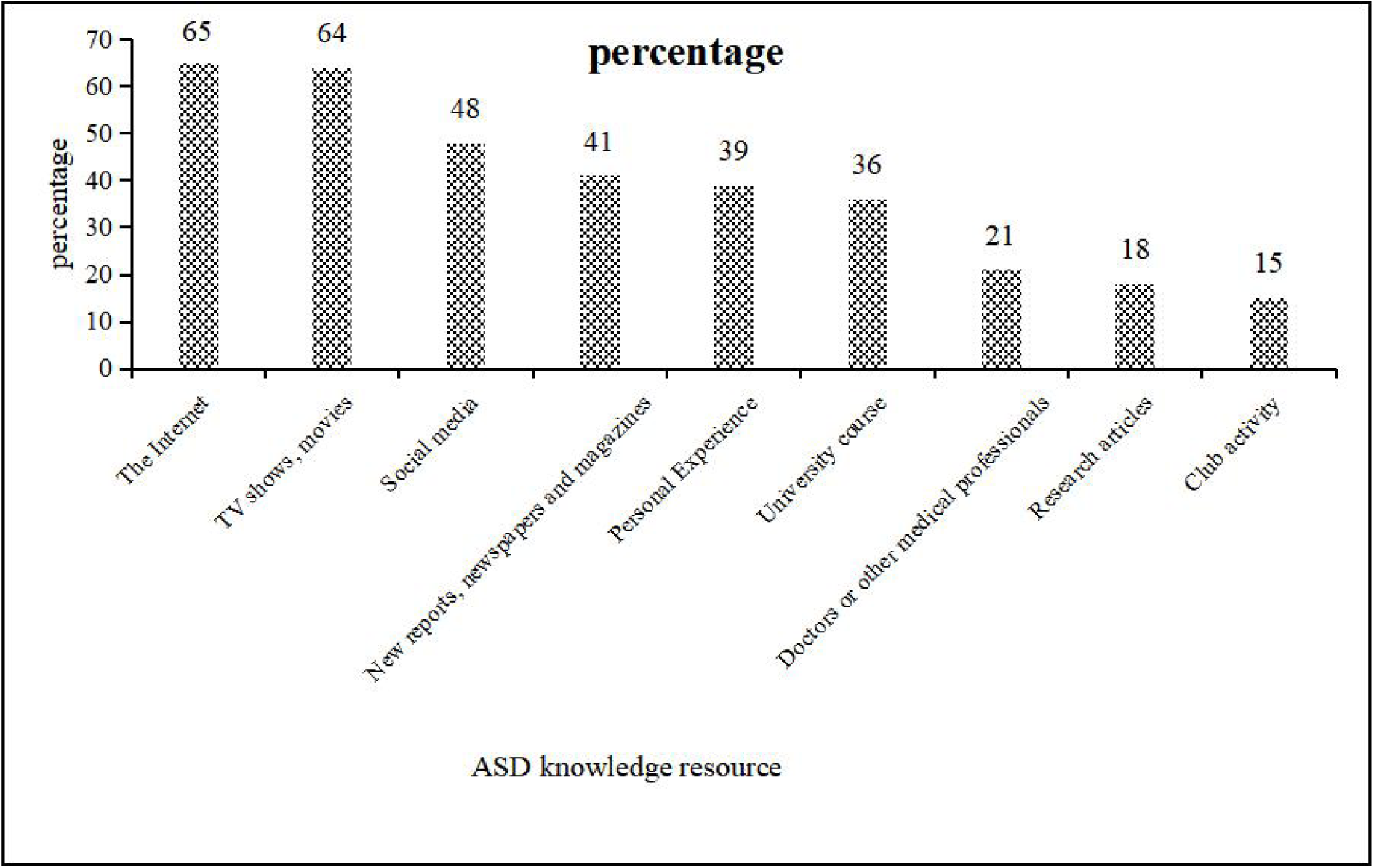
Percentage of students’ ways of acquiring ASD knowledge resources.

## Discussion

### Analysis of various demographic factors

Gender has consistently been a significant factor, with numerous studies indicating that females possess greater ASD knowledge and lower stigma endorsement compared to males [7][36–39].

In this study, grade level also emerged as a variable with significant differences, with senior students demonstrating the highest level of ASD knowledge and the lowest stigma endorsement. Higher grade levels correspond to increased knowledge acquisition throughout university education, reflecting a higher level of education. Previous literature has consistently shown a strong correlation between education level and ASD knowledge, with more highly educated participants exhibiting greater ASD knowledge and more positive attitudes towards autisitic people [36–38][40,41].

University students who have interacted with autistic people have a deeper understanding of ASD and lower stigma endorsement. This finding aligns with conclusions from previous research. For instance, Lan (2020) found that 244 normal-education students showed explicit cognitive stigma and low-intensity implicit stigma towards autistic children; contact experiences promote more positive explicit attitudes among normal-education students towards autistic children[39]. However, it’s worth noting that some studies have shown that interacting with autistic people isn’t necessarily a positive experience. Huskin’s research (2018) pointed out that frequent contact with autistic children may actually reduce the willingness to interact [42].When people have limited knowledge or misconceptions about ASD, they tend to exhibit idealized positive social attitudes[43]. Yet, after having had some experience with autistic people and having some knowledge of them, they may have lower levels of acceptance and openness [44]. Future research should delve into how different types and qualities of contact (such as frequency, depth, and context) influence an individual’s knowledge and attitudes towards ASD, examining the distinct mechanisms by which positive and negative contacts affect stigma, to facilitate the design of effective educational interventions.

### Factor analysis of different academic backgrounds

In this study, we observed that normal-education students exhibited greater knowledge of ASD and lower stigma endorsement compared to non-normal-education students. Particularly, students majoring in special education demonstrated superior knowledge of ASD and lower stigma. This may be attributed to their systematic professional training, which includes understanding, identifying, and intervention strategies for ASD. Such accumulated expertise enables them to comprehend the needs of autistic students more accurately, thereby reducing misconceptions and stigma towards this group. Students who have taken inclusive education courses, although not majoring in special education, also received diverse inclusive values in their curriculum, including correct recognition and acceptance of children with special needs, which similarly helps to diminish misunderstandings and prejudices towards this group. Additionally, normal-education students have more opportunities to interact with special needs students during their coursework and internships, which enhances their understanding of ASD and reduces biases. This study supports the recommendation of incorporating special education courses into normal-education students programs and suggests that inclusive education courses can enhance university students’ awareness of special education, providing a theoretical basis for the future popularization of such courses in non-normal-education students programs.

### Improve the dissemination of ASD knowledge and education strategies

The survey results align with prior research, highlighting the Internet and social media as key platforms for university students to access information and communicate [36]. However, the uneven quality of these channels may foster misunderstandings or stereotypes[45,46]. Future research should prioritize optimizing dissemination channels to promote accurate and authoritative ASD knowledge. Additionally, university courses and medical expertise are underrepresented, underscoring the need to integrate ASD-related content into curricula, encourage research engagement, and support community activities to enhance theoretical and practical knowledge. Introducing professional resources such as doctors and psychologists can further provide credible, high-quality knowledge. These measures can help students develop a comprehensive understanding of ASD, reduce stigma, and foster an inclusive society.

## Data Availability

All data produced in the present study are available upon reasonable request to the authors.

## Acknowledgments

None.

## Supporting information

S1 Table1.

S2 Fig1.

